# Sorghum utilization in grain-based food products in China and Australia

**DOI:** 10.1101/2025.10.21.25338131

**Authors:** Anita Stefoska-Needham, Sophie L. Marsano, Liyan Zhong, Thomas H. Roberts

## Abstract

Sorghum consumption has potential health-promoting effects for consumers. This study identified and compared sorghum-containing grain-based food products available in major supermarkets in China and Australia. A total of 1,692 products were audited in Shenzhen, China and Illawarra, Australia, in 2023/24. Breakfast cereals, snack bars, flours, pastas, and noodles were evaluated. Information on ingredients, including the presence of sorghum, food format, brand, product name, wholegrain/gluten-free labelling was recorded. In China, sorghum was found in 4.3% (12/279) of breakfast cereals, with only 1/12 sorghum-containing breakfast cereals listed sorghum in the first position of the ingredient list. Sorghum was found in 2.0% (9/458) of snack bars and was listed as either ‘sorghum’ (n=3) or ‘sorghum flour’ (n=6). In Australia, sorghum was found in 22/356 (6.2%) breakfast cereals, 9/285 (3.2%) snack bars, and was absent from all flours, pastas, and noodles. Most sorghum-containing cereals were extruded (36.4%) and labelled gluten-free (16/22, 73%) or wholegrain (14/22, 64%). Sorghum-containing snack bars, notably oat-bake and muesli bars, were mostly made from sorghum flour and flakes. Sorghum appeared in the first position in the ingredient list in 2/22 (9.1%) of breakfast cereals, and in the third or higher position for all snack bars. Among the breakfast cereal and snack bar subcategories analyzed, there were no significant differences in sorghum utilization between China and Australia (Fisher’s Exact Tests, p < 0.05), except for oat bake snack bars (higher in China, p = 0.0265). Overall, the audit data supported the conclusion that sorghum is underutilized as an ingredient in common grain-based food products available to consumers in major Chinese and Australian supermarkets. Greater awareness of its potential consumer health benefits is needed to drive utilization of sorghum grain in foods across different markets.

## 1. Introduction

Sorghum (*Sorghum bicolor* (L.) Moench) is a gluten-free, wholegrain cereal crop cultivated globally. In regions of the world such as north-west and south-central Africa, sorghum grain is traditionally consumed in staple foods, whereas in Western countries it is typically used in animal feed for domestic use or export, and to a much lesser extent in human foods [1]. The ability to grow sorghum in rain/temperature-variable climates, even in regions experiencing significant water scarcity, exemplifies sorghum’s potential to contribute to sustainable agriculture and food security around the world [2].

Nutritionally, sorghum is important due to its association with disease-mitigating mechanisms, notably those underpinning the development of cardiovascular disease, diabetes mellitus and some types of cancer [3, 4]. The current evidence base, as appraised in a recent systematic literature review [5], indicates improvements in markers of oxidative stress, control of blood glucose and lipid levels, and enhancement of satiety/appetite sensations associated with weight management, particularly when wholegrain sorghum foods are consumed regularly. Experimental animal and cell-line research also indicates evidence for favourable cell-mediated immune responses, including antioxidant and anti-inflammatory effects, particularly for polyphenol-rich red and brown sorghum grain varieties [6].

Regular consumption of sorghum grain is needed for consumers to gain the purported health benefits of this cereal. However, this can be difficult to achieve in societies where sorghum is not consumed traditionally, such as in Australia, and the sorghum-containing product range is limited [7, 8]. Given that sorghum grain—like its counterparts: wheat, rye, barley and oats—is most likely to be used in the production of cereal-based foods (breakfast cereals and bakery goods), a limited range of sorghum products on supermarket shelves may be a barrier to enabling the potential consumer health benefits.

In countries with a limited range of sorghum-containing food products, more investment in food innovation is required to increase the volume and variety of sorghum-containing food products available to consumers. This must be done with consideration of consumers’ expectations of what constitutes healthy cereal-based foods. Consumer research indicates that the perceived healthiness of cereal foods is associated with greater wholegrain, dietary fibre, and antioxidant contents, as well as lower levels of salt and fat [9], which is likely to drive consumer purchases. From a nutrition science perspective, the beneficial, synergistic effects of the germ and bran fractions of wholegrains are typically implicated in health benefits associated with cereals, given they are high in dietary fibre, healthy lipids, micronutrients, and phytochemicals [10-12]. Sorghum food products can also be manufactured to possess these characteristics, and hence to be marketed as ‘healthy’, as well as ‘gluten-free’; however, attention must be given to the impact of processing methods on the health-enhancing potential and sensory appeal of end-products [13, 14].

Monitoring progress towards greater sorghum-based product expansion and food innovation in Western societies requires regular evaluation of the prevalence of sorghum in products available to consumers globally. A world-first (to our knowledge) study of sorghum prevalence was conducted in 2020 through a cross-sectional audit of Australian supermarkets [15]. Sorghum ingredients were found in only 6.1% of ready-to-eat breakfast cereals and 2.0% of snack bars. The audit was limited to these two food categories in major supermarkets, and excluded smaller, speciality food retail outlets, posing a limitation given that innovations, with lesser-known ingredients such as sorghum, are more likely to be sold in such stores [15]. This study provided important baseline data, and the authors recommended expansion to other categories in future audits to measure the prevalence in a wider range of food products.

China, a major global economic market, consumes huge amounts of sorghum grain, but it is mostly used to make Baijiu, the dominant Chinese white spirit. Nevertheless, sorghum grain is reported to be present in an expanding range of food products in Chinese supermarkets, including bread and noodles [16], although this is yet to be substantiated. Investigating the applications of sorghum in Chinese food products could offer valuable insights into how sorghum grain is utilized in this market and help drive demand for sorghum as a human food by consumers, supported by sorghum growers and the food industry globally. Hence, the present study was conducted to examine the prevalence of sorghum in a range of grain-based food products in a cross-section of supermarkets in China and Australia. A secondary aim was to compare trends observed in the present audits to the previously mentioned, seminal Australian audit conducted in 2020.

## 2. Materials and methods

### 2.1 Location of study and supermarket selection

The China-based audit was conducted in the city of Shenzhen in Guangdong Province, southern China, in three major Chinese supermarkets—Bravo, Vanguard, and Walmart—in 2023. Larger supermarkets located near central Shenzhen were selected to ensure the widest range of food products on offer. The Australia-based audit was conducted in the Illawarra region of New South Wales in 2024. Retail outlets were selected to represent a range of socio-economic status (SES) and market share (Woolworths 38.2%, Coles 29.0%, other companies 17.6% and Aldi 8.6%), maximizing access to a greater variety of products. Woolworths, Coles and Aldi supermarkets in Unanderra (low SES), Wollongong (medium SES) and Figtree (high SES) were approached for inclusion in the audit. Targeted health food/speciality stores were in Wollongong, but only one store agreed to participate. Permission was received from store managers prior to conducting the audits in both China and Australia.

### 2.2 Product selection

Product categories were selected on the basis that they contained grain-based foods. In China, breakfast cereals and snack bars were selected for analysis, as per the 2020 audit conducted in Australia [15]. In the present Australian-based audit, breakfast cereals and snack bars were also included, as well as flour, pasta and noodle products.

### 2.3 Data collection

In China, data were collected over one week in February 2023. Eligible products were located in the conventional breakfast cereal and snack bar sections of all three supermarkets. The speciality sugar-free health food sections (commonly found in Chinese supermarkets) were also scanned for breakfast cereal products. In Australia, data collection took place over 3 weeks in April 2024. Eligible products were located in designated supermarket aisles for each category. Breakfast and snack bar products in the health food aisles were also assessed, given that grain-based food products increasingly dominate these locations in Australian supermarkets.

Data collection involved photographing the front-of-package, nutrition information panel (NIP), and ingredients list of each product. Only products in the largest package size were recorded to avoid duplication, and, if a product was sold at multiple store locations, it was counted only once. As per the methods reported previously [15], the following details were recorded for each product according to its product category: brand with product name, store location, product type (specific to each category), and presence or absence of sorghum in the ingredient list. The ingredients were only recorded for products in which sorghum was identified, including sorghum’s position in the ingredients list. In both China and Australia, ingredients must be listed in descending order by in-going weight, meaning the first ingredient listed contributes to the largest weight in the product and the last ingredient contributes to the least weight. The following product details were also recorded: percentage contribution of sorghum (if available), sorghum format (e.g. wholegrain, flaked or puffed), any health/ nutrition related claims or food attributes (if present, e.g., gluten free), target audience (if stated, e.g., children). In the Australian audit, a note was made if the same product was included in the 2020 audit.

### 2.4 Data analysis

Descriptive statistics were used to analyze the data using Microsoft Excel 365 (version 16.50). Counts and percentages were used to evaluate the prevalence of sorghum in different product categories (breakfast cereals, snack bars in both China and Australia; flours, pastas and noodles in Australia only) and their subcategories, which were based on sorghum format, e.g., flaked or puffed. Where relevant, observations of trends compared to the 2020 Australian audit results were made [15].

To enable a statistical comparison between sorghum prevalence in supermarkets in China and Australia, various product subcategories were combined to reduce the total number of subcategories. The counts of sorghum-containing products were too low to allow valid Chi-square tests to be performed; therefore, a Fisher’s Exact Test was performed on each contingency table (2×2), using GraphPad Prism v10.5.0. A Fisher’s Exact Test was also performed on the total breakfast cereal counts and total snack bar counts, again comparing China to Australia.

## 3. Results

In this study, a total of 1,692 products were evaluated across five different grain-based product categories in China and Australia. In China, 737 food product items were analyzed. Of these, 279 items were categorized as breakfast cereals, while the remaining 458 items were categorized as snacks. In Australia, a total of 955 products were audited, and of these, 356 were breakfast cereals, 285 were snack bars, 80 were flours, 59 were noodles, and the remaining 175 products were pastas.

### 3.1 Breakfast cereal audit in China

Porridge-style cereals were most frequently identified in the China-based sample of products (93/279, 33.3%), followed by bubbles/puffs/flakes (27.2%), and muesli (24.7%). Of the 279 cereals examined, only 12 (4.3%) contained sorghum across a range of breakfast cereal types, including porridge (n=5), bubbles, flakes and puffs (n=3), muesli (n=3), and flaked biscuits (n=1) (Table 1).

**Table 1.**
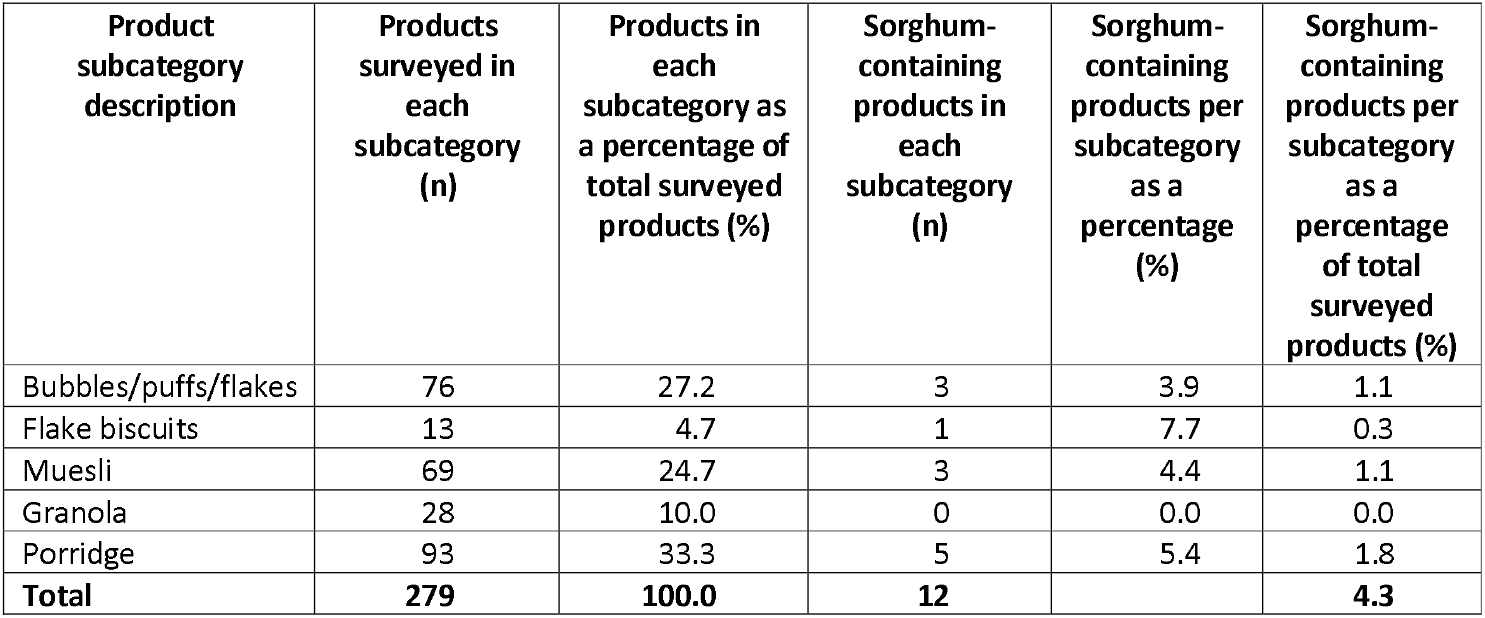
Summary of sorghum-containing breakfast cereals in three major supermarkets in Shenzhen, China. Data are presented as counts (n) and percentages (%).

| Product subcategory description | Products surveyed in each subcategory (n) | Products in each subcategory as a percentage of total surveyed products (%) | Sorghum-containing products in each subcategory (n) | Sorghum-containing products per subcategory as a percentage (%) | Sorghum-containing products per subcategory as a percentage of total surveyed products (%) |
| --- | --- | --- | --- | --- | --- |
| Bubbles/puffs/flakes | 76 | 27.2 | 3 | 3.9 | 1.1 |
| Flake biscuits | 13 | 4.7 | 1 | 7.7 | 0.3 |
| Muesli | 69 | 24.7 | 3 | 4.4 | 1.1 |
| Granola | 28 | 10.0 | 0 | 0.0 | 0.0 |
| Porridge | 93 | 33.3 | 5 | 5.4 | 1.8 |
| **Total** | **279** | **100.0** | **12** |  | **4.3** |

Of the sorghum-containing breakfast cereals identified, only 1/12 listed sorghum in the first position of the ingredient list. Sorghum was listed second in the ingredient list in 2/12 of the products, and in the third position or beyond in 9/12 products. The most common sorghum ingredient in the ingredient list was sorghum flour (5/12), followed by wholegrain sorghum (3/12), sorghum (3/12), and organic sorghum (1/12). Eight of the 12 (66.7%) sorghum-containing breakfast cereal products had wholegrain sorghum listed as an ingredient, while the remaining products did not specify whether the grain was wholegrain. Among the breakfast cereal subcategories, sorghum was most frequently used in flake biscuits and bubbles/puffs/flakes: 7.7% (1/13) and 3.9% (3/76), respectively (Table 1).

A total of (7/12) sorghum-containing breakfast cereals were gluten-free, and 3/12 were marketed towards children and were made with wholegrain sorghum. Most of the sorghum-containing breakfast cereals (9/12) were in the health food section of the supermarket. Only 3/12 of the sorghum-containing breakfast cereals mentioned ‘sorghum’ on the front-of-pack label, with the remainder mentioning ‘sorghum’ only in the list of ingredients or in small font on the back of the pack. All 12 sorghum-containing breakfast cereals included additional descriptors such as ‘organic sorghum’, ‘sorghum rice’, ‘sorghum flour’, or ‘puffed sorghum’.

### 3.2 Breakfast cereal audit in Australia

Breakfast cereals audited in Australia comprised 12 subcategories: biscuit, bran sticks, cluster, combination cereal products, extruded shapes, filled, flakes, granola, muesli, oats, porridge and puffs/bubbles (Table 2). Of these subcategories, granola was the most common (51/356, 14.3%), followed by muesli (47/365, 13.2%) and porridge (45/365, 12.6%). Of the total cereals assessed, only 22 of 356 (6.2%) contained sorghum as an ingredient. The top six formats of sorghum ingredients were extruded shaped cereals, followed by biscuit, granola muesli, porridge, combination cereal products and flakes. Sorghum was not identified in any breakfast cereal products in the speciality health food store audited.

**Table 2.** Summary of sorghum-containing breakfast cereals in major supermarkets and a smaller health food store in the Illawarra region of New South Wales, Australia. Data are presented as counts (n) and percentages (%).

| Product subcategory description | Products surveyed in each subcategory (n) | Products in each subcategory as a percentage of total surveyed products (%) | Sorghum-containing products in each subcategory (n) | Sorghum-containing products per subcategory as a percentage (%) | Sorghum-containing products per subcategory as a percentage of total surveyed products (%) |
| --- | --- | --- | --- | --- | --- |
| Biscuit | 17 | 4.8 | 5 | 29.4 | 1.4 |
| Bran sticks | 5 | 1.4 | 0 | 0.0 | 0.0 |
| Cluster | 39 | 11.0 | 0 | 0.0 | 0.0 |
| Combination cereal products | 21 | 5.9 | 1 | 4.8 | 0.3 |
| Extruded shapes | 39 | 11.0 | 8 | 20.5 | 2.2 |
| Filled | 1 | 0.3 | 0 | 0.0 | 0.0 |
| Flakes | 38 | 10.7 | 1 | 2.6 | 0.3 |
| Granola | 51 | 14.3 | 3 | 5.9 | 0.8 |
| Muesli | 47 | 13.2 | 2 | 4.3 | 0.6 |
| Oats | 34 | 9.5 | 0 | 0.0 | 0.0 |
| Porridge | 45 | 12.6 | 2 | 4.4 | 0.6 |
| Puffs/bubbles | 19 | 5.3 | 0 | 0.0 | 0.0 |
| **Total** | **356** | **100.0** | **22** |  | **6.2** |

Sorghum was found in extruded-shaped breakfast cereals more commonly (36.36%) compared to breakfast cereal subcategories (Table 2). Sorghum was positioned first in the ingredients list in 2/22 (9.1%) of sorghum-containing breakfast cereal products, listed second in 7/22 (31.8%) of products, third in 10/22 (45.5%) of products and greater than third in 3/22 (13.6%) of products. In these breakfast cereal products, sorghum was most commonly listed as sorghum flour (n=13), followed by sorghum (n=3), wholegrain sorghum flour, and sorghum.

Sixteen of the 22 products (73%) were gluten-free, two of which stated ‘gluten-free flour blend’ in the ingredients list, but further details relating to the type of sorghum flour were not provided. Children were the target market for 5/22 (23%) of these products, based on label designs including images. Only 4/22 (18%) of products spelled out the word ‘sorghum’ anywhere on the packaging outside of the ingredients list. Wholegrain claims were found on 14/22 (64%) of breakfast cereal products, and out of these products, 6/22 (27%) contained wholegrain sorghum in the ingredient list. All the products that contained wholegrain sorghum in the ingredients list made wholegrain claims. ‘Source of fibre’ was a popular claim with all products making a fibre statement on the front-of-pack label.

### 3.3. Snack bar audit in China

A total of 458 snack products were analyzed in eight subcategories (Table 3). Only 2.0% (9/458) of the snack bar products contained sorghum as an ingredient. Of these, three were classified as biscuit bars (33%), two as muesli/cereal bars (22%), two as puff/bubble bars (22%), one as oat bake bars (11%), and one as a rolled-oat bar (11%).

**Table 3.** Summary of sorghum-containing snack bars in three major supermarkets in Shenzhen, China. Data are presented as counts (n) and percentages (%).

| Product description/style | Products surveyed in each subcategory (n) | Products in each subcategory as a percentage of total surveyed products (%) | Sorghum-containing products in each subcategory (n) | Sorghum-containing products per subcategory expressed as a percentage (%) | Sorghum-containing products per subcategory as a percentage of total surveyed products (%) |
| --- | --- | --- | --- | --- | --- |
| Biscuit | 108 | 23.6 | 3 | 2.8 | 0.7 |
| Cake | 72 | 15.7 | 0 | 0.0 | 0.0 |
| Muesli | 58 | 12.7 | 2 | 3.4 | 0.4 |
| Nut/seed | 63 | 13.8 | 0 | 0.0 | 0.0 |
| Oat bake | 32 | 7.0 | 1 | 3.1 | 0.2 |
| Pressed | 28 | 6.1 | 0 | 0.0 | 0.0 |
| Puff/bubble | 12 | 2.6 | 2 | 16.7 | 0.4 |
| Rolled oat | 85 | 18.6 | 1 | 1.2 | 0.2 |
| **Total** | **458** | **100.0** | **9** |  | **2.0** |

The utilization of sorghum varied across the different types of snack bars. In biscuit-style bars, muesli/cereal-style bars, and oat bake-style bars, sorghum was incorporated in formulations as ‘sorghum flour’ or ‘wholegrain sorghum’. Sorghum was identified in the 8^th^ or higher position in the ingredients list. However, in the other categories, ‘puffed sorghum’ was listed in the ≥5^th^ positions in the ingredients list, and the product descriptor included ‘puffed product’.

The specification of whether wholegrain sorghum ingredients were used in the bars was not provided by the food manufacturers. All sorghum bars were labelled gluten-free, with 3/9 (33%) marketed toward infants and children. Although none of the snack bars mentioned sorghum in their product names, some displayed sorghum on the front-of-pack labels. Additionally, none of the sorghum-containing snack bars included the term ‘ancient grain(s)’ in their ingredient lists across all subcategories.

### 3.4 Snack bar audit in Australia

In Australia, the snack bar audit comprised 285 products across 15 subcategories (Table 2). These were biscuit style, cake bar, cereal bar, filled, muesli bar, nut bar, oat bake bar, oat bar, popcorn bar, pressed bar, protein bar, protein muesli bar, protein nut bar, puff/bubble, seed bar. Of these categories, nut bars were the most common (21.4%), followed by pressed bars (20.7%) and oat bake bars (10.9%). Of the audited bars, nine out of 285 (3.2%) contained sorghum as an ingredient (Table 4). Sorghum was not identified in any snack bar products in the speciality health food store audited.

**Table 4.** Summary of sorghum-containing snack bars in major supermarkets and a smaller health food store in the Illawarra region of New South Wales, Australia. Data are presented as counts (n) and percentages (%).

| Product subcategory description/style | Products surveyed in each subcategory (n) | Products in each subcategory as a percentage of total surveyed products (%) | Sorghum-containing products in each subcategory (n) | Sorghum-containing products per subcategory as a percentage (%) | Sorghum-containing products per subcategory as a percentage of total surveyed products (%) |
| --- | --- | --- | --- | --- | --- |
| Biscuit | 1 | 0.4 | 0 | 0.0 | 0.0 |
| Cake | 7 | 2.5 | 0 | 0.0 | 0.0 |
| Cereal bar | 10 | 3.5 | 0 | 0.0 | 0.0 |
| Filled | 19 | 6.7 | 0 | 0.0 | 0.0 |
| Muesli | 26 | 9.1 | 2 | 7.7 | 0.7 |
| Nut | 61 | 21.4 | 0 | 0.0 | 0.0 |
| Oat bake | 31 | 10.9 | 7 | 22.6 | 2.5 |
| Oat | 18 | 6.3 | 0 | 0.0 | 0.0 |
| Popcorn | 1 | 0.4 | 0 | 0.0 | 0.0 |
| Pressed | 59 | 20.7 | 0 | 0.0 | 0.0 |
| Protein | 14 | 4.9 | 0 | 0.0 | 0.0 |
| Protein muesli | 11 | 3.8 | 0 | 0.0 | 0.0 |
| Protein nut | 7 | 2.5 | 0 | 0.0 | 0.0 |
| Puff/ bubble | 17 | 5.9 | 0 | 0.0 | 0.0 |
| Seed | 3 | 1.0 | 0 | 0.0 | 0.0 |
| **Total** | **285** | **100.0** | **9** |  | **3.2** |

The sorghum-containing snack bars comprised only two categories, oat bake bars (n=7) and muesli bars (n=2). Sorghum’s position in the ingredients list was greater than position three for all the snack bar products audited. Sorghum flour (n=7) and sorghum flakes (n=6) were the most common formats in the snack bars, followed by puffed sorghum (n=2). No bars specifically identified that the sorghum utilized was wholegrain. Of the sorghum-containing snack bars audited, none were perceived to be specifically targeted at children. All the sorghum-containing snack bars were gluten-free. Two out of the nine (22%) sorghum-containing snack bars stated the word “sorghum” on the product packaging, as well as in the ingredients list.

### 3.5 Audit of pastas, noodles, and flour products in Australia

The audit of pastas, noodles, and flour products did not identify sorghum in any ingredient formulations, including generically labelled gluten-free options in these product categories.

### 3.6 Comparison of sorghum prevalence in breakfast cereals and snack bars between supermarkets in China and Australia

Among the breakfast cereal (Table 5) and snack bar (Table 6) subcategories analyzed, there were no significant differences in sorghum utilization between China and Australia (Fisher’s Exact Tests, p< 0.05), except for oat bake snack bars, of which one (of 31) were from China and seven (of 24) from Australia (p = 0.0265).

**Table 5.** Fisher’s Exact Tests of sorghum prevalence in breakfast cereals in supermarkets in China versus Australia. Data are derived from Tables 1 and 2.

| Product subcategory description/style | Country | Sorghum-containing products in each subcategory | Non-sorghum-containing products in each subcategory | P value |
| --- | --- | --- | --- | --- |
| Bubbles/puffs/flakes | China | 3 | 73 | 0.6349 |
|  | Australia | 1 | 56 |  |
| Biscuits/flake biscuits | China | 1 | 12 | 0.1961 |
|  | Australia | 5 | 12 |  |
| Muesli/granola | China | 3 | 94 | 0.7208 |
|  | Australia | 5 | 93 |  |
| Porridge | China | 5 | 88 | >0.9999 |
|  | Australia | 2 | 43 |  |
| All others | China | 0 | 0 | >0.9999 |
|  | Australia | 9 | 130 |  |
| **Total** | China | **12** | **267** | 0.3751 |
|  | Australia | **22** | **334** |  |

**Table 6.**
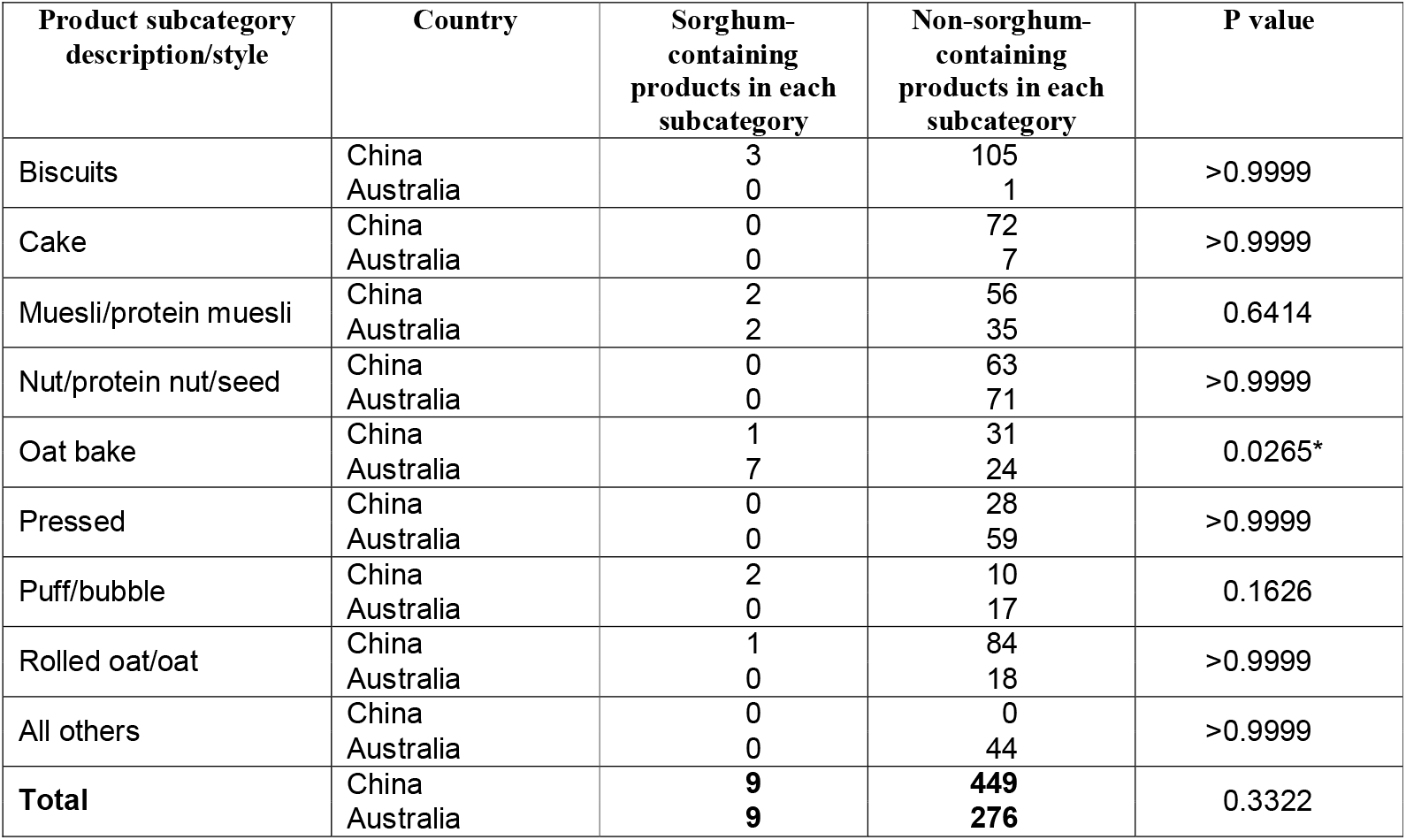
Fisher’s Exact Tests of sorghum prevalence in snack bars in supermarkets in China versus Australia. Data are derived from Tables 3 and 4. Contingency tables are 2 × 2. An asterisk highlights p values < 0.05.

## 4. Discussion

In this cross-sectional study, we investigated the utilization of sorghum in grain-based food products across food retail outlets in China and Australia. To our knowledge, this study represents the first benchmarking of sorghum’s prevalence in cereal-based food products in China, and builds upon prior research in Australia that included a seminal audit conducted in 2020 [15]. Overall, sorghum prevalence in Chinese breakfast cereals and snacks was similar to that observed in Australia, with sorghum identified in 8.3% of 279 breakfast cereals and 2.0% of 458 snack bars audited in China, compared to 6.2% of 356 breakfast cereals and 3.2% of 285 snack bars in Australia. The Australian findings represent a slight rise from 6.1% for breakfast cereals and 2.0% for snack bars in 2020 [15]; however, sorghum was absent from all pastas, noodles and flour products reviewed in the new Australian audit.

In China, approximately one-third of sorghum-containing breakfast cereals were identified in the porridge subcategory, unlike in Australia, where sorghum was more commonly found in ready-to-eat extruded breakfast cereals. In China, porridge is often prepared using a powdered or paste base that is mixed with milk or water, offering a novel format with potential applicability in Australian markets. In these mixtures, sorghum is typically combined with other grains such as corn, barley, and red bean to create a porridge-type product. Whether this type of product has potential as a breakfast cereal alternative for Australian consumers requires testing and validation by risk-tolerant food product developers/manufacturers interested in new product launches.

The breakfast cereal markets in both China and Australia have experienced significant growth due to increasing consumer interest in healthy and nourishing ready-to-eat breakfast options. From 2012 to 2017, the market in China showed a yearly growth rate of 10.2% [17], and it is projected to reach a revenue of US$1.32 billion in 2024 [18]. This high demand presents an opportunity for commercial success in the breakfast cereal market by expanding the sorghum breakfast cereal product range in both countries. Targeting young people aged between 2 and 18 years old may be a key strategy for sorghum food innovators and research and development, given that 44.1% of all wholegrain products annually are consumed by this group in Australia [15, 19]. In China, grain foods are staples in the diets of consumers across all age groups and typically include rice and wheat products (such as noodles and steamed buns), but the prevalence of sorghum in these commonly consumed foods is currently unknown. Noodles and pasta products were reviewed in the Australian audit presented in this paper, but sorghum was not identified in any formulations.

Compared with breakfast cereals, sorghum prevalence in snack bars was observed to be lower in both China and Australia, likely due to the significantly smaller snack bar market relative to the breakfast cereal market in these countries [20, 21]. Snack bar formulations in China were largely comprised of sorghum flour, which is produced by milling sorghum grain to formulate different types of snack bar formats, including the more popular biscuit-style and rolled oat-style bars. These applications appear to be an extension of traditional Chinese foods, such as mantou, offering potential ideas for novel product innovation in this category in Australia.

Although sorghum flour was identified in the snack bar products in China, it was rarely the dominant or characterizing ingredient. In the biscuit-style snack bars, it typically comprised about 5% of the ingredients, alongside other grains like oats (≥ 5%) and corn (≥ 5%). In the Australian audit, nut bars were observed to still dominate since 2020, potentially explaining the low prevalence of sorghum in this category [15]. Like in China, the sample of sorghum-containing snack bars comprised mostly sorghum flour, followed by sorghum flakes then puffed sorghum. This contrasted with the 2020 audit, where sorghum flour was not listed in the ingredients list of any snack bars in Australia, and puffed sorghum was most common [15].

The marketing of sorghum in both breakfast cereals and snack bars is limited in both countries. For example, in Australia, only two of the snack bars included ‘sorghum’ on the packaging, and it was listed for its texture contribution rather than its health effects. The inclusion of sorghum on the front-of-packaging would be a step in the right direction to increase consumers’ awareness of sorghum, as lack of recognition has been identified as a major barrier to acceptance of sorghum as a human food [7].

In China, the healthier sorghum-containing snack bars, characterized by higher contents of wholegrain and/or fibre and low or no added sugars, tended to be marketed towards children and younger people. Examples of these products included wholegrain sticks and brown rice crackers. In contrast, in Australia, there were no sorghum-containing snack bar products targeted towards children, despite being a convenient and easy lunch box option. According to the earlier Australian audit [15], manufacturers of the grain-based snack bars may have little motivation to explore new formulation ideas, such as using sorghum alongside more fibre and less sugar, due to their strong market position with current offerings. However, demonstrating other food successes in other markets, such as in China, may stimulate ideas for new product development to increase market share in Australia. Furthermore, wholegrain sorghum was not listed in the ingredients of any snack bars containing sorghum in Australia, nor was there a wholegrain claim displayed, despite wholegrain claims generally being used more frequently on Australian food products [22]. Research by the Grains and Legumes Nutrition Council of Australia indicates that Australian consumers do not prioritize consumption of wholegrains, despite having an awareness of their potential health benefits [23], implying that marketers may not be interested in leveraging wholegrain claims to drive consumer purchases in the snack bar product category.

A cross-section of speciality health food retailers was intended to be included in the Australian audit presented here; however, only one store agreed to be involved. Despite this, important insights were afforded by this single site. Although sorghum was not found in any breakfast cereals, snack bars, flours, pastas or noodles, the diversity of product offerings reflected more specialized, health-focused products, particularly in the breakfast cereal and flour categories. These findings suggest potential for sorghum’s incorporation into novel formats. Expanding future audits to include more health food and speciality shops could provide a more comprehensive understanding of emerging trends. Including gym and fitness stores, which cater to health-conscious consumers, may further reveal opportunities for sorghum innovation. Also, future audits should focus on ethnic food speciality stores, especially to capture people from cultural backgrounds that use sorghum in traditional cooking, such as people from African and Indian communities. The bread category should also be audited in future across a variety of retail outlets, including supermarkets, speciality food stores and bakeries, as bread is the most frequently purchased grain food in Australia after refined pasta and rice [23].

Future opportunities for sorghum food innovation in both China and Australia should leverage the potential health benefits and food attributes associated with wholegrains broadly. Consumer preferences in both countries are shifting towards health-conscious and diet-focused choices, with many consumers willing to pay a premium for perceived health benefits [24, 25]. The rising demand for gluten-free products has also opened opportunities for smaller producers, driving innovation and competition [26]. The healthy breakfast and snack food market will likely grow in response to increased demand for convenient, health-oriented products. Wholegrain foods, particularly those with a low glycemic index, have also gained popularity [27], presenting opportunities for expanding sorghum-based offerings. The growing interest in plant-based diets [28] also positions sorghum as a viable ingredient for meeting these evolving demands, potentially increasing its acceptance and prevalence in human food supplies. Targeting young consumers (up to 18 years) in both countries presents an opportunity to expand the sorghum breakfast and snack bar cereal market, as this demographic contributes significantly to wholegrain consumption [19]. In China, sorghum is most recognized for its use in liquor production; however, during the auditing of products for this study, sorghum was also identified in cooking sauces and snacks (results not reported). This highlights sorghum’s versatility and potential for diverse applications [16].

The key strengths of this study include the rigorous methodology and cross-country analysis, which provided valuable insights into sorghum’s market presence. The large sample size and standardized data collection methods strengthen the study’s reliability. However, between-country differences in food labelling regulations, dietary preferences, and the inability to specify sorghum content due to labelling practices constrained direct comparisons. Limited access to health food and speciality shops in Australia also restricted the scope of the audit. Ethical considerations prevented identifying specific brands or stores, and ingredient percentage data for sorghum was not always available, limiting the granularity of the analysis.

## 5. Conclusions

This study highlights a positive trend in sorghum’s integration into grain-based food products in China and Australia and identifies opportunities for further market penetration through innovation and education. Emphasizing sorghum’s health benefits and expanding its applications in novel and traditional formats can foster greater consumer acceptance and demand. Strategic product development, combined with targeted marketing focused on health benefits, can enhance sorghum’s appeal to consumers, fostering greater adoption in diverse food categories.

Future research should explore the nutritional and sensory benefits of sorghum-containing products and assess their appeal across diverse consumer groups. Expanding audits to additional food categories and evaluating new product formats could uncover further opportunities for sorghum innovation. Audits of speciality retailers, including those selling diverse ethnic foods as well as health food stores, would also provide a more comprehensive understanding of emerging trends and opportunities.

## Data Availability

All relevant data are within the manuscript.

## Authorship contributions

**Anita Stefoska-Needham**: Conceptualization; Research design; Data analysis; Original manuscript draft; Review and editing; Academic supervision of Liyan Zhong and Sophie L. Marsino. **Sophie L. Marsino**: Data collection; Data analysis; Writing initial results for the Australian audit under the academic guidance of Anita Stefoska-Needham. **Liyan Zhong**: Data collection; Data analysis; Writing initial results for the Chinese audit under the academic guidance of Thomas H. Roberts. **Thomas H. Roberts**: Conceptualization; Research design; Data analysis; Original manuscript draft; Review and editing; Academic supervision of Liyan Zhong. All authors have read and agreed to the published version of the manuscript.

## Acknowledgments

We acknowledge Cecily Ducksbury for guidance to Sophie L. Marsino in data collection. We thank Associate Professor Floris van Ogtrop (University of Sydney) for statistical advice.

## Data Availability

All relevant data are within the manuscript. Data linking specific products with individual supermarkets are commercially sensitive and thus cannot be shared publicly.

